# Efficiency of Artificial Intelligence in Detecting COVID-19 Pneumonia and Other Pneumonia Causes by Quantum Fourier Transform Method

**DOI:** 10.1101/2020.12.29.20248900

**Authors:** Erdi Acar, Bilge Öztoprak, Mustafa Reşorlu, Murat Daş, İhsan Yılmaz, İbrahim Öztoprak

**Affiliations:** Dept. of Computer Engineering, Institute of Science, Çanakkale Onsekiz Mart University, Çanakkale, Turkey; Dept. of Radiology, Faculty of Medicine, Çanakkale Onsekiz Mart University, Çanakkale, Turkey

**Keywords:** COVID-19, Pneumonia, Deep Learning, CT Denoising, Quantum Fourier Transform

## Abstract

The new coronavirus (COVID-19) appeared in Wuhan in December 2019 and has been announced as a pandemic by the World Health Organization (WHO). Currently, this deadly pandemic has caused more than 1 million deaths worldwide. Therefore, it is essential to detect positive cases as early as possible to prevent the further spread of this outbreak. Currently, the most widely used COVID-19 detection technique is a real-time reverse transcription-polymerase chain reaction (RT-PCR). However, RT-PCR is time-consuming to confirm infection in the patient. Because RT-PCR is less sensitive, it provides high false-negative results. Computed tomography (CT) is recommended as a solution to this problem by healthcare professionals because of its higher sensitivity for early and rapid diagnosis. In addition, radiation used in CT poses a serious threat to patients. In this study, we propose a CNN-based method to distinguish COVID-19 pneumonia from other types of viral and bacterial pneumonia using low-dose CT images to reduce the radiation dose used in CT. In our study, we used a data set consisting of 7717 CT images of 350 patients that we collected from Çanakkale Onsekiz Mart University Research Hospital. We used a CNN-based network that suppresses noise to remove interference from low-dose CT images. In the image preprocessing phase, we provided lung segmentation from CT images and applied quantum Fourier transform. By evaluating all possible variations of local knowledge at the same time with quantum Fourier transformation, the most informative spatial information was extracted. In CNN-based architecture, we used pre-trained ResNet50v2 as a feature extractor and fine-tune by training with our dataset. We visualized the efficiency of the ResNet50v2 network using the t-SNE method. We performed the classification process with a fully connected layer. We created a heat map using the GradCam technique to see where the model focuses on the images while classifying. In this experimental study, the results of 99.5%, 99.2%, 99.0%, 99.7%, and 99.1%, were obtained in the context of performance criteria such as accuracy, precision, sensitivity, specificity, and f1 score, respectively. This study revealed the artificial intelligence-based computer-aided diagnosis (CAD)system as an effective and fast method to accurately diagnose COVID-19 pneumonia.

## 1 Introduction

COVID-19 is a contagious disease caused by the mutant SARS-CoV-2 virus with high mortality[1]. After the description of the first case in Wuhan, China in December 2019, it rapidly spread all over the world and announced to be a pandemic by WHO on 11 March 2020. It causes a disease with a wide range of symptoms: Mainly fever, cough, headache, weakness, sore throat, loss of taste and smell, diarrhea, chest pain, shortness of breath, feeling pressure on the chest, conjunctivitis, skin eruptions, and discoloring in distal parts of extremities. Most of the patients who tested positive for COVID-19 are either asymptomatic or have a mild disease characterized by fever and dry cough. Elderly with chronic underlying diseases are more prone to develop a serious disease. Mortality is mostly due to multiorgan failure caused by the cytokine storm. Computed tomography (CT) is widely used for the detection of lung lesions in highly suspected patients. It is also used in the course of the disease to detect the extent of lung involvement and complications. COVID-19 pneumonia is characterized by patchy areas of ground-glass opacity (GGO) in both lungs predilecting peripheral areas of posterior and lower segments. In most advanced cases interlobular septal thickening accompanies GGOs leading to the typical “crazy paving” pattern. Other findings include consolidation, dilatation of vessels in the affected areas of lungs, subpleural bands, bronchiectasis, hyperlucent areas in GGOs-the so-called vacuolar sign, air bronchograms, pleural and pericardial effusion, and enlarged lymph nodes[2]. As one would expect, none of these findings is specific for COVID-19 pneumonia and there is considerable overlap between COVID-19 pneumonia and other infectious and noninfectious lung diseases. Typical peripheral multifocal ground-glass appearance allow sitting to be differentiated from bacterial pneumonia. However, it is difficult to distinguish from other types of viral pneumonia and atypical pneumonia because of findings that coincide [3,4].

There have always been attempts to differentiate between diseases in radiology since the vast majority of lesions on radiological images are nonspecific and it is sometimes impossible to differentiate between different diseases manifesting as similar radiological abnormalities.

Currently, the reverse transcription real-time fluorescent polymerase chain reaction (RT-PCR) test based on nucleic acid amplification test (NAAT) of respiratory tractor blood samples is used to diagnose a patient with symptoms of COVID-19[5]. Nevertheless, its sensitivity and detection rate are low due to the low viral load early. So, it is likely to give inaccurate results. In addition, it may take time to yield the test result. The progression and severity of the infection cannot be estimated since it gives only positive or negative results [6].

Chest radiography may be normal in early/mild disease. Thorax CT is more sensitive than a chest x-ray for early COVID-19 diagnosis[7]. If a patient with suspected COVID-19 has negative test results but findings are present on CT imaging, the patient should be quarantined, and the RT-PCR test is repeated. This rapid isolation is crucial to proper patient care and disease control. Because it provides fast and accurate results, CT plays an important role in the early diagnosis of COVID-19 pneumonia. However, to obtain high-quality images, a high radiation dose must be administered to patients. This entails a potential risk of genetic damage and cancer related to the radiation, thus, finding a solution to minimize radiation is crucial. One approach to reducing the risk of radiation is to use lower levels of x-ray current, but quantum noise caused by an insufficient number of photons increases the noise and artifacts in low dose computed tomography (LDCT) images. This could potentially impair diagnostic performance. Asa solution to this problem, many methods have been designed for denoising LDCT images. These methods generally fall into three categories: techniques based on sinogram filtering, iterative reconstruction, and post-reconstruction image processing[8–10]. In addition, denoising algorithms using artificial deep neural networks have demonstrated impressive performance capabilities in this context[8–17].

Recently, with the advancement of artificial intelligence technology and especially the development of convolutional neural networks, computer vision has led to extraordinary developments. It is widely used in the medical field and provides support for medical diagnosis. Additionally, due to the heavy workload on a large number of infected patients and healthcare professionals, the artificial intelligence-based computer-aided system can speed up the diagnostic process. These systems can provide an efficient diagnosis of the disease in a shorter time, such as the lack and inability of radiologists during the pandemic crisis. It can also reduce the need for human surveillance and identify details invisible to the human eye. Artificial intelligence (AI) technologies are rapidly developing in the area of medicine, specifically in radiology, to overcome this challenge[18]. In this context, many deep learning models have also been developed for the diagnosis of COVID-19[19–33].

The aim of this study is to test the effectiveness of AI distinguishing COVID-19 pneumonia from other types of pneumonia by using low-dose CT images to reduce the radiation used in CT. The paper is organized as follows. In section 2, material and method are given. In section 3, obtained results are given. In section 4, discussion and concluding remarks are given with comparing the other studies in the literature.

## 2 Material and Method

In this study, we used a data set consisting of 7717 CT images of 350 patients obtained from Çanakkale Onsekiz Mart University Research Hospital. We used an LDCT noise removal deep model that removes noise from LDCT images. By modifying the Bi-Directional ConvLSTM U-Net[34] architecture, we provided lung segmentation with a deep neural network. We applied quantum Fourier transform[35] at the image pre-processing stage. We used ResNet50v2[36] architecture as a feature extractor. We visualized the efficiency of the feature extractor using t-Distributed Stochastic Neighbor Embedding (t-SNE)[37] on the resulting feature vector. We used in-place / on-the-fly data augmentation[38] during training to increase the quality and robustness of the model instead of increasing the number of data in the data set. We used the Gradcam[39] technique to visualize where the model focuses on the images while classifying the images. We use a full connected layer for classification. We evaluated performance results with accuracy, precision, sensitivity, specificity, and f1-score.

### 2.1 Dataset

The dataset was collected from Çanakkale Onsekiz Mart University Research Hospital with ethical permissions. The dataset includes 1157 Bacterial pneumonia, 4387 COVID-19, 695 Viral pneumonia, and 2635 normal diagnosed 7717 lung CT images in total. The images belong to 350 patients taken on different days during the past one year and the images belonging to the COVID-19 class belong to patients with positive RT-PCR tests. Each of the images has a standard of 512×512 pixel size. An example of the dataset is given in Fig. 1. In addition, the distribution of images in the dataset is visualized in Fig. 2 using t-SNE which is a size reduction technique that is well suited for visualizing high dimensional datasets.

**Fig. 1.**
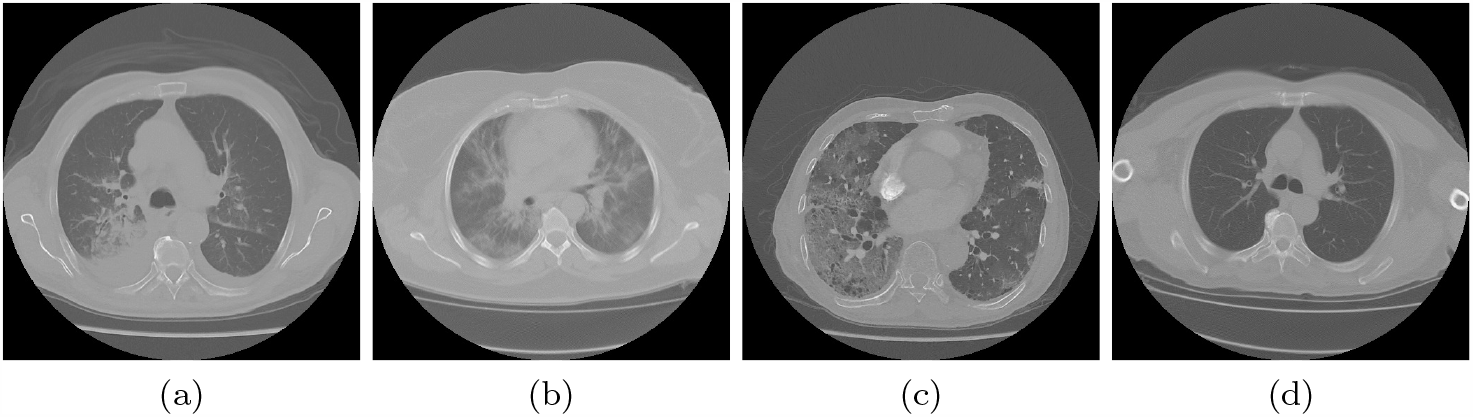
Sample images of classes within the dataset. **a** is an example of Bacterial Pneumonia.**b** is an example of COVID-19. **c** is an example of Viral Pneumonia. **d** is an example of Normal.

**Fig. 2.**
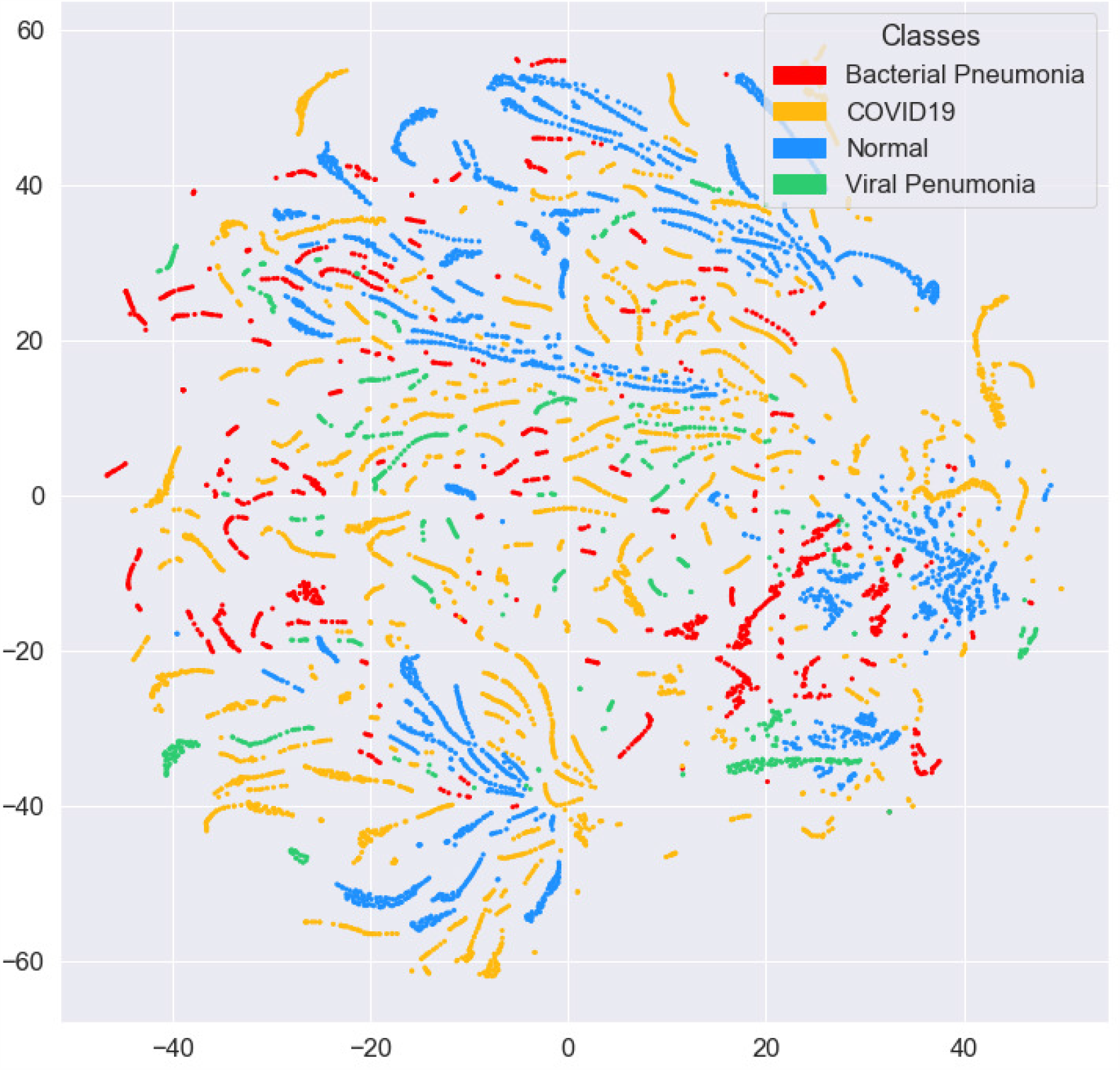
Distribution of the dataset visualized using t-SNE.

### 2.2 Low Dose CT Simulation

The performance of deep learning models has always been debatable due to the insufficiency of educational samples in medical datasets. A deep learning model learns the distribution of all the training data it sees during the training process. If there is not enough data to train the network, an over-fitting problem will occur. Low-dose and normal-dose CT data are needed to train the low-dose CT noise suppression network. However, it is not always possible to find such a dataset. Therefore, we created the low dose CT dataset using the normal dose CT images we had. The noise that occurs in CT images due to insufficient number of photons in CT images taken by reducing radiation is ‘quantum noise’. It is also known to has Poisson noise and a Poisson distribution[40]. We applied the procedure of Zeng et al.[41] to obtain LDCT image. We obtained sinogram data by applying Radon transformation to NDCT images and added Poisson noise to that sinogram data [42]. Then, we obtained LDCT images by applying inverse radon transform to the Poisson noise-added sinogram data. LDCT sample images are given in Fig. 3.

**Fig. 3.**
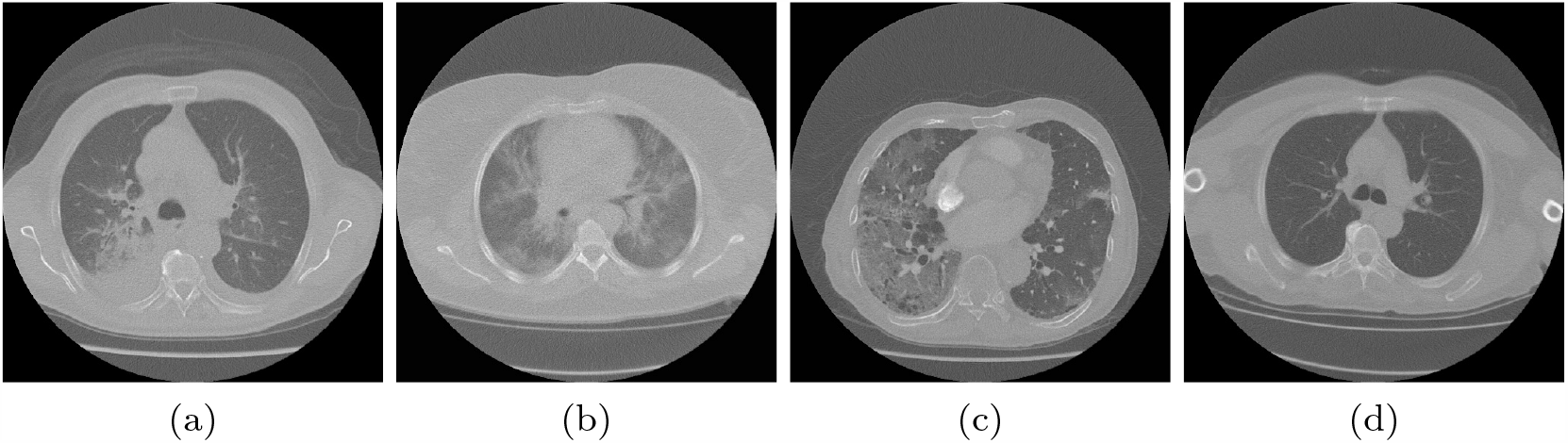
LDCT sample images of classes within the dataset. **a** is an example of low dose bacterial pneumonia.**b** is an example of low dose COVID-19. **c** is an example of low dose viral pneumonia. **d** is an example of low dose normal.

### 2.3 Low Dose CT Denoising Architecture

We applied a deep residual neural network (ResNet) for image-to-image transformation in an attempt to predict full-dose CT from low-dose CT images. We modified the model proposed by Yang et al.[44] to remove noise. This network consists of two parts: (1) Denoising network. (2) Perceptual loss calculator. Fig. 4 presents the architecture of denoising employed in the current study. To learn denoising images containing different structures and densities, a sufficiently deep network to handle complexity is required. Some studies have indicated that networks with more layers do not always perform well, and that training loss is increasing. As a solution to this problem, He et al.[36] proposed a residual learning model. In our work, we applied residual learning between the lower and upper layers. As shown in Figure 4, the input image and output of layers 1, 2 and 3 are combined with the output of layers 7, 6 and 5, respectively. These links pass the details of the image to lower layers because the properties on the first layers contain more input information.

**Fig. 4.**
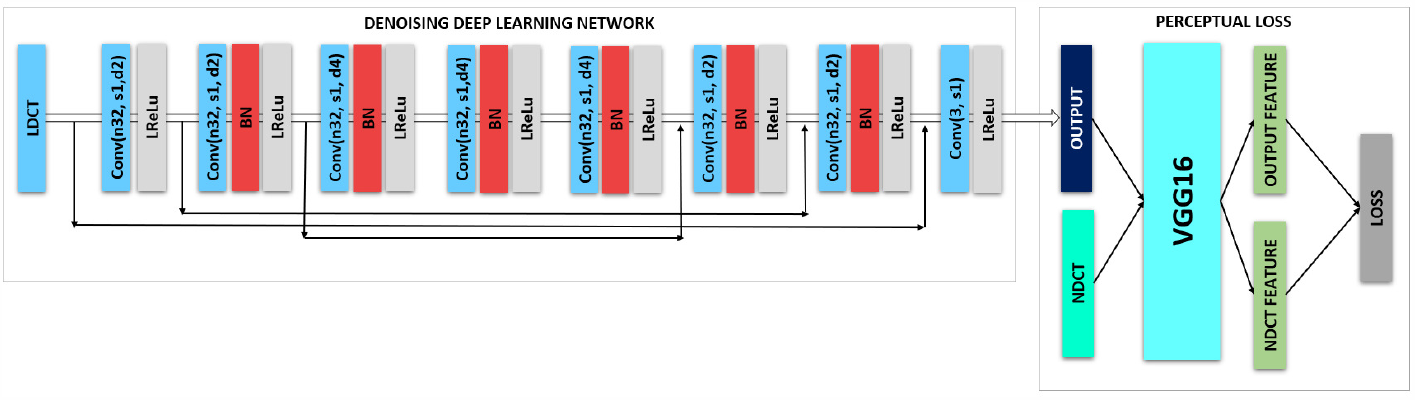
LDCT denoising deep learning network architecture

In our study, the denoising network consists of 8 convolutional layers. We used 32 filters with 3×3 kernel size in the first 7 layers. We applied 3 convolutional filters with 3×3 kernel size in the layer that is the output of the network only. We applied batch normalization(BN) in 6 layers to speed up the training process. We used Leak Rectified Linear Unit (LReLU) as the non-linear activation function for the 8 convolutional layers. Also, we use dilation rate in each layer to increase receptive field. The receptive field (RF) which is the region of the input image used to calculate the output value. A larger receptive field means more contextual information is captured than the input image.

Perceptual loss is calculated in the second part of the proposed network using the VGG16 CNN architecture[43]. We started the VGG16 network with imagenet weights, then retrained and fine-tuned the dataset. The output of the noise denoising network is fed as input to the VGG16 network and normal dose CT images are provided as labels. Loss is calculated by using the features from the network and back propagation is performed.

We used 80% of the dataset for training and 20% of the dataset for testing the noise removal network. We used the Adam stochastic gradient algorithm to optimize the model. The learning rate was started at 1e-3 and dynamically adjusted. Structural similarity index (SSIM) and peak signal-to-noise ratio (PSNR) measurements were used in the quantitative evaluation of the predicted NDCT images. Evaluation results are given in Table. 1.

**Table 1.** Quantitative evaluation results for LDCT denoising network.

| Train Size | Test Size | SSIM | PSNR |
| --- | --- | --- | --- |
| 6174 | 1543 | 0.9726 | 36.95 |

### 2.4 Lung Segmentation

Since distinctive information of pneumonia is in the lungs on CT images, we apply lung segmentation to obtain only the lung area. For segmentation, we applied a similar approach to BidirectionalConvLSTMU-Net architecture. The architecture of the segmentation network is given in Fig. 5. We used 70% of the dataset for training and 30% of the data set for testing in the training of the segmentation network. We initially received the 1e-4 learning rate and dynamically reduced it. We trained using the Adam stochastic optimization algorithm which was developed as a solution to the vanish gradient problem. The performance results of the segmentation network are given in Table. 2.

**Table 2.** Performance results for segmentation network.

| Train Size | Test Size | Accuracy | Error Rate | S-Dice |
| --- | --- | --- | --- | --- |
| 5401 | 2316 | 0.9870 | 0.0325 | 0.9934 |

**Fig. 5.**
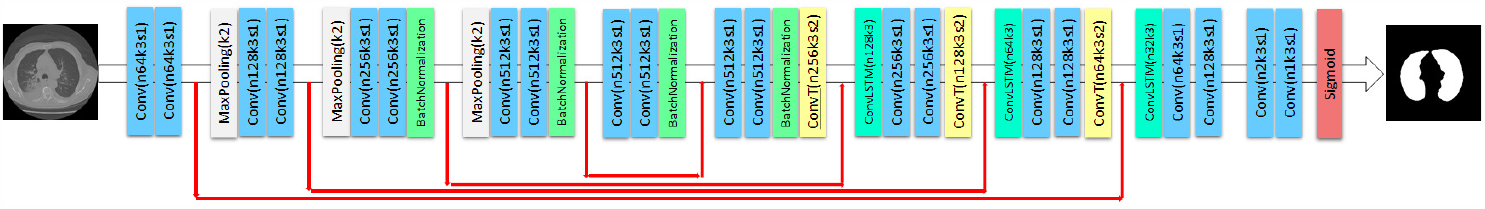
The architecture of the segmentation network.

We obtained an accuracy rate of 0.9870, error rate of 0.0325 and s-dice rate of 0.9934. The region of interest is obtained from the CT images using the Graphcut image processing method. Sample images of the lung regions obtained after the graphcut procedure are given in Fig. 6.

**Fig. 6.**
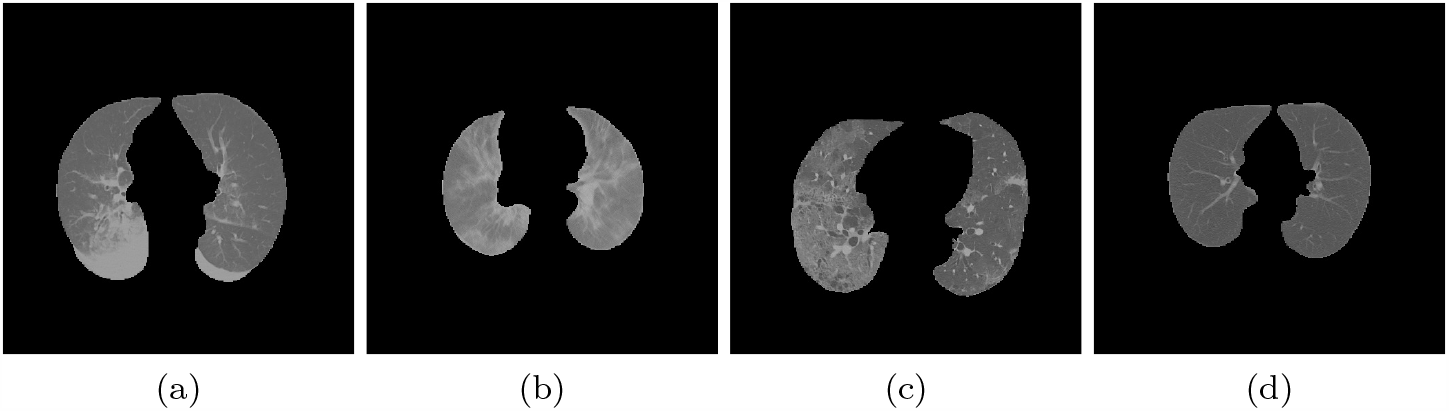
Sample lung regions obtained as a result of segmentation from the images in the data set. **a** is an example of lung region for bacterial pneumonia.**b** is an example of lung region for COVID-19. **c** is an example of lung region for viral pneumonia. **d** is an example of lung region for normal.

Then all images in the dataset are brought to 256×256 pixels in size and the intensity pixel values of all images are normalized from [0,255] to the standard normal distribution by min-max normalization to the intensity range of [0,1] as follows.

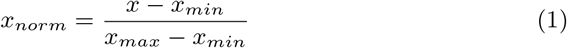

where *x* is the pixel intensity. *x*_*min*_ and *x*_*max*_ are minimum and maximum intensity values of the input image.

### 2.5 Data Augmentation

The performance of deep learning neural networks often increases with the amount of data available. Data augmentation is a technique of artificially generating new training data from existing training data. Image data augmentation involves creating transformed versions of images in the training dataset that belong to the same class as the original image. Conversions involve a number of operations from the image manipulation area such as pan, rotate, zoom, and much more. The aim is to increase the generalizability of the model while expanding the training data set with new, reasonable examples.

Image data augmentation is applied to the training dataset. Given that network is constantly seeing new, slightly modified versions of input data, the network can learn more robust features. There are three types of data augmentation methods: (1) Dataset generation and expanding an existing dataset. The problem with this approach is that the generalization capability of the model may not fully increase. (2) In-place/on-the-fly data augmentation. Using this type of data augmentation, it is ensured that the network is trained to see new variations of data in each epoch. This method increases the generalization ability of the model. (3) A hybrid approach combining 1 and 2 approaches. We use an in-place / on-the-fly augmentation approach when training the network. In addition, we used flip left-right (probability = 0.3) and rotate (probability = 0.4, max left rotation = 10, max right rotation = 10) augmentation techniques.

### 2.6 Quantum Fourier Transform

Pattern recognition is currently applied in many fields such as medicine, space, defense industry, security and sensor technologies. These processes take a long time in conventional computers due to the increasing number of data. It is emphasized that such problems can be solved more easily with quantum computers that are still developing. In addition to such a situation, quantum machine learning can improve machine learning performance and make it efficient due to quantum’s superior properties such as entanglement and superposition. In the context of quantum machine learning, relevant research is carried out on whether quantum information brings a new perspective to how machines recognize patterns in data, whether they can learn from less training data, their ability to develop new machine learning methods, or to identify[45,46].

There are two different strategies when designing quantum machine algorithms. The first is to translate classical machine learning models into quantum computing language to achieve an algorithmic acceleration. The second is to combine classical algorithms with quantum tools, keeping computational resources low. In our study, we adopt the second approach and apply quantum fourier transform (QFT) in the image preprocessing stage. QFT circuit is given in Fig. 7.

**Fig. 7.**
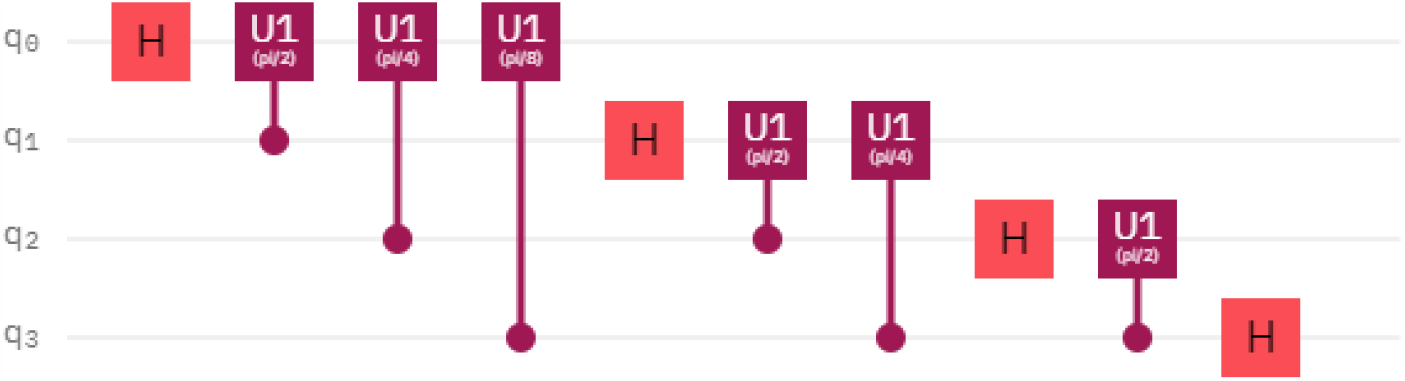
Quantum fourier transform circuit.

With QFT, any quantum state is moved into phase space. Thus, all possible situations are easily recognized and calculated simultaneously with the effect of the superposition state and semantically valuable information is extracted. Thus, QFT can be used as a feature extractor.

For applying QFT, we first encode the image into a quantum circuit. QFT application scheme is given in Fig. 8. We use a 4-qubit quantum circuit for the 2×2 kernel filter used in QFT. The quantum circuit we use consists of encoding data, applying the quantum fourier transform, and a measurement process called decoding. During the data coding phase, the Hadamard gate is applied to each qubits and all qubits are brought into superposition. The input data is then encoded into the quantum circuit as parameters of the Ry gates. Then, CNOT gates are applied between 1-2, 2-4 and 2-3 qubits so that each qubit becomes entangled with each other and the data encoding phase is completed. In the second step, the QFT circuit is applied. In the decoding stage, the measurement process is performed to obtain the expected value of each qubit on the basis of Pauli-Z. The measurement results are written into the feature matrix.

**Fig. 8.**
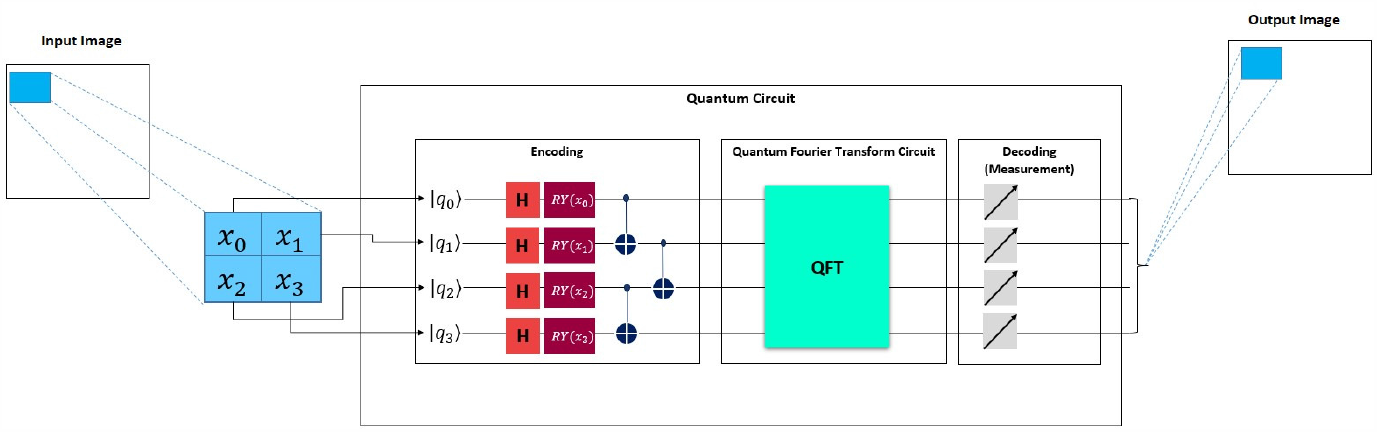
Quantum fourier transform implementation scheme.

### 2.7 Framework of Proposed Deep Learning Model For Diagnostic

In the model, we used the ResNet50v2 network as a feature extractor. We used 2×2 kernel size in the QFT. In addition, we started the ResNet50v2 network with imagenet weights. Then we applied the flatten layer, which converted the feature matrix obtained from the feature extractor into a feature vector so that we could feed it into the full connected layer. Then we applied dense layers (512, ReLu), Batch Normalization (0.2), (256, ReLu), and (4, Softmax). We used the NDCT dataset formed as a result of the output of the LDCT denoising network in the training of the network. The architecture of the proposed model is given in Fig. 9.

**Fig. 9.**
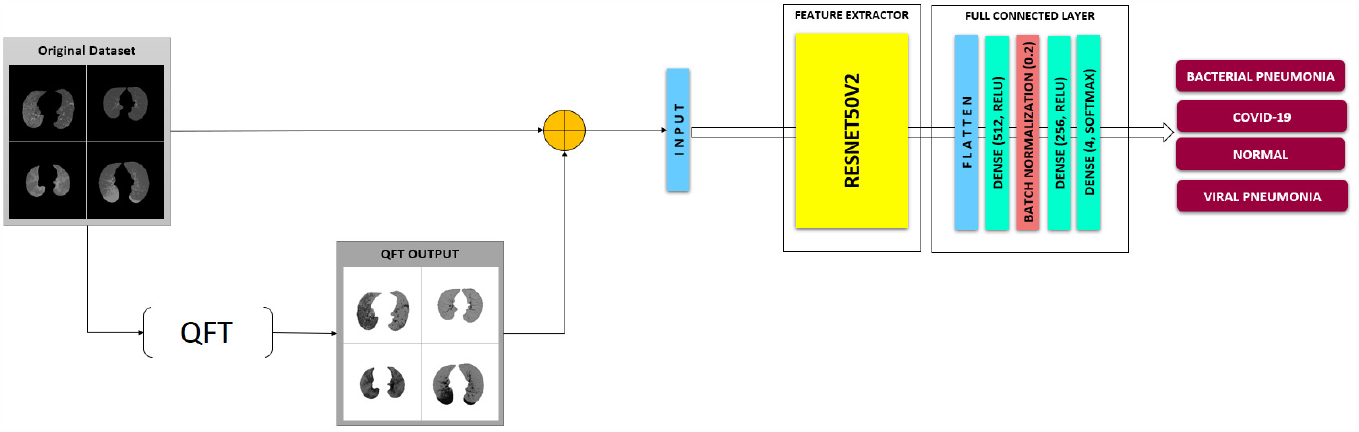
Framework of proposed deep learning network for diagnostic.

## 3 Result

### 3.1 Performance Metrics

We reported the performance of the proposed model with accuracy, precision, sensitivity, specificity and F1-score metrics. The confusion matrix used to explain the performance of the classification model consists of true positive (TP), true negative (TN), false positive (FP), and false negatives (FN).

Accuracy is a measure of how many correct predictions the classifier makes of all test data and is defined as follows.

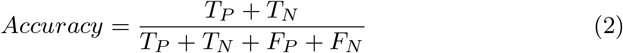

Precision is an indication of how many of the positive classes are positively predicted r and is calculated as follows.

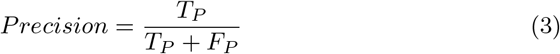

Sensitivity refers to how well anticipates those who are actually sick and is calculated as follows.

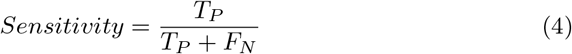

Specificity is the proportion of people who test negatively among those who actually do not have the disease and is defined as:

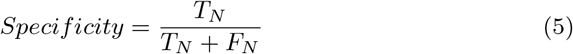

Precision and sensitivity are two important measures and there is a trade-off between them. F1-Score is a good choice when you want to deal with this trade-off and strike a balance between them. This score can be interpreted as a weighted sensitivity and precision average and is defined as follows.

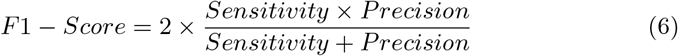

### 3.2 Experimental Results and Analysis

The dataset used in the training of the proposed model consists of NDCT images obtained in the output of the LDCT denoising network.We reserve %80 of the dataset for training and %20 of the dataset for testing. The number of data allocated for training, validation and testing is given in Table. 3. To speed up the training phase, the images are scaled and the network takes a 256 x 256-pixel image as input. We optimize the network with the Adam optimization algorithm. We start the learning rate at 0.001 and dynamically reduce it during training. Besides, we apply early stopping when no increase is observed in the training process.

**Table 3.** Dataset distribution used for training and test.

| Classes | Train Size 72% | Validation Size 8% | Test Size 20% |
| --- | --- | --- | --- |
| Bacterial Pneumonia | 832 | 93 | 232 |
| COVID-19 | 2326 | 258 | 646 |
| Viral Pneumonia | 500 | 56 | 139 |
| Normal | 1898 | 210 | 527 |
| Total | 5556 | 617 | 1544 |

The architecture of the quantum fourier transform in the preprocessing stage was run on the qiskit quantum simulator. For the rest of the model, we’re using the TensorFlow interface. The study was carried out on a personal computer with an Intel Xeon E3-1245 3.7 GHz processor, 48 GB RAM, and 2 GB Nvidia P400 graphics card.

The change of the training and validation phase of the proposed deep learning model by epoch is given in Fig. 10. Loss denoted how the actual outcome differs from the estimate. The loss is expected to be inversely proportional to the accuracy of the model. Considering that the weights were randomly selected at the beginning of the training from the graph in Fig. 10, it showed low accuracy and high loss values as an indication that the network made false predictions. However, the accuracy value showed higher values in the following periods, and the error rate decreased. This indicated that the model was beginning to solve the classification problem. We are started training 30 epochs in model training. After the 15th epoch, the model reached the highest accuracy value and the lowest loss value. After this epoch, it was seen that the performance did not improve.

**Fig. 10.**
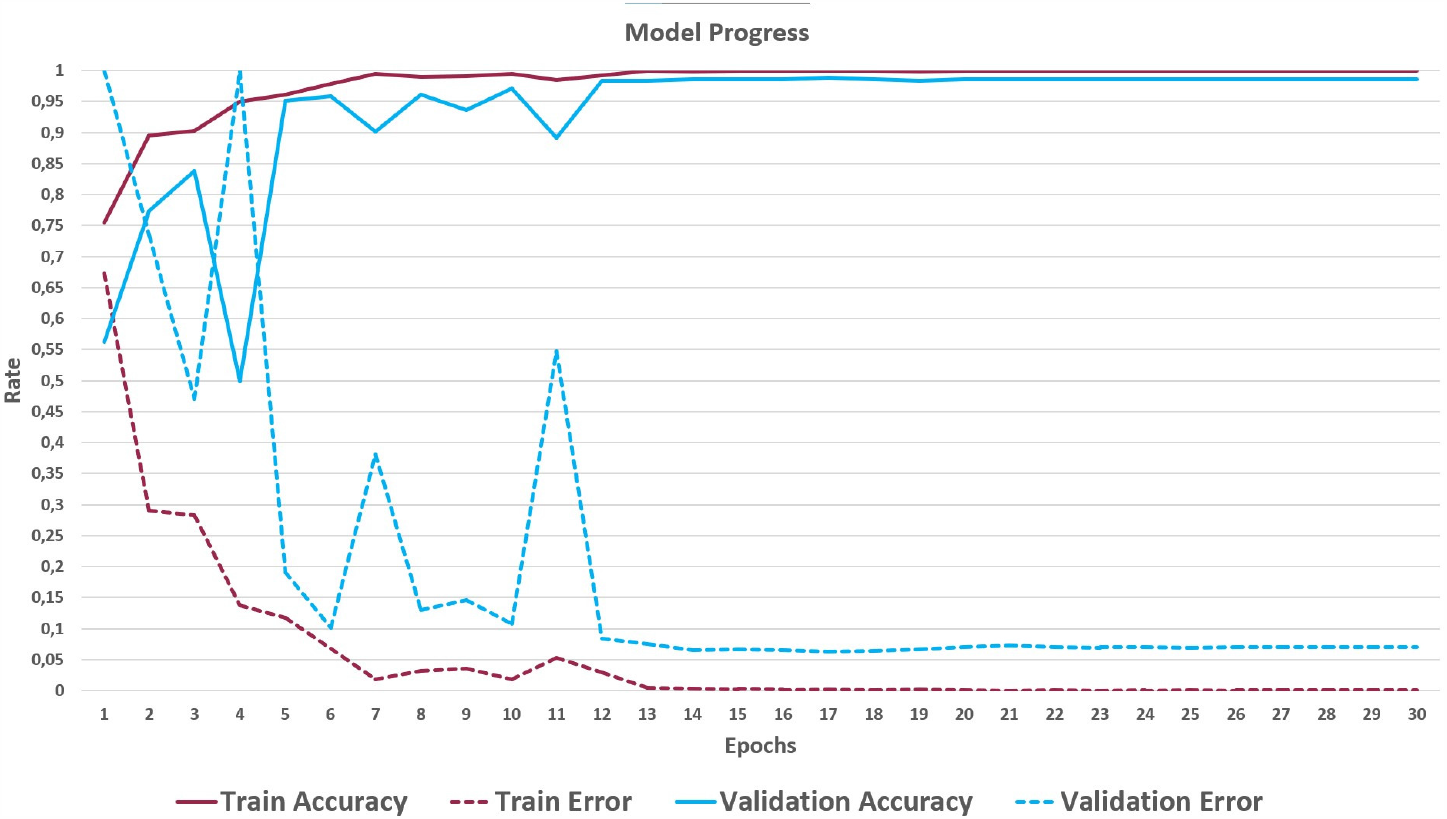
The change of the training and validation phase of the proposed deep learning model by epoch.

The dataset used to show the efficiency of the ResNet50v2 feature extraction network and its visualization using the test dataset t-SNE are given in Fig. 12. Considering Fig.2 and Fig.12 comparatively, it can be observed that ResNet50v2 effectively extracts different features of the classes in the dataset in which it performs effectively.

**Fig. 11.**
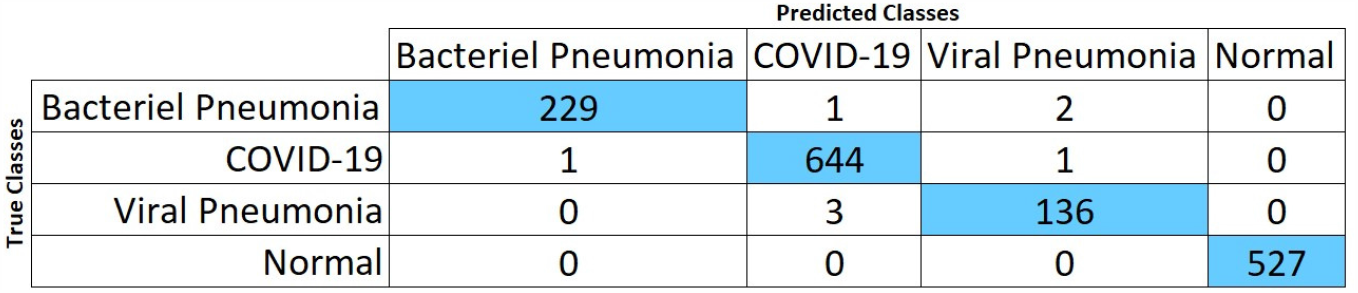
Confusion matrix values of the proposed model.

**Fig. 12.**
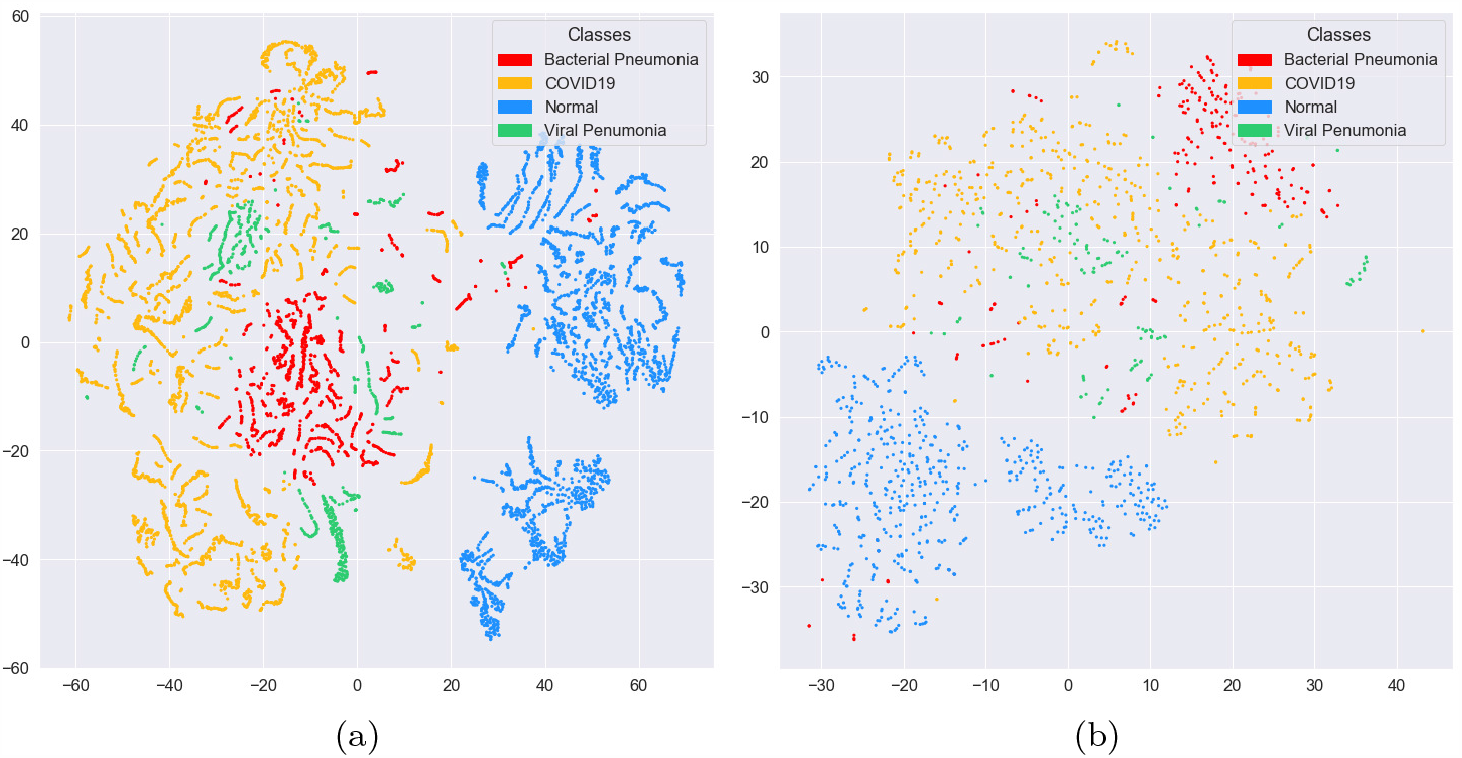
**(a)** is distribution of data set after feature extraction visualized using t-SNE. **(b)** is distribution of test data set after feature extraction visualized using t-SNE.

The confusion matrix and performance results of the proposed model is given in Fig. 11 and Table. 4 respectively. The overall performance results of the proposed model are 99.5%, 99.2%, 99.0% 99.7%, 99.1% for accuracy, precision, sensitivity, specificity, and F1-score, respectively.

**Table 4.** The results show performance results for the proposed model.

| Accuracy: | 0.995 |  |  |  |
| --- | --- | --- | --- | --- |
| Error Rate: | 0.021 |  |  |  |
|  | Precision | Sensitivity | Specificity | F1-Score |
| Bacterial Pneumonia | 0.996 | 0.987 | 0.999 | 0.991 |
| COVID-19 | 0.994 | 0.997 | 0.991 | 0.995 |
| Viral Pneumonia | 0.979 | 0.979 | 0.998 | 0.979 |
| Normal | 1.000 | 1.000 | 1.000 | 1.000 |
| Overall | 0.992 | 0.990 | 0.997 | 0.991 |

To increase the interpretability of the model, we used the gradient-weighted class activation mapping (GradCAM) method to visualize where the model focuses on the CT images while making decisions. We created heat maps with this mapping method. This method shows suspicious areas in COVID-19 and other pneumonia images. Sample images for the heat map of COVID-19 and other pneumonia images are given in Fig. 13

**Fig. 13.**
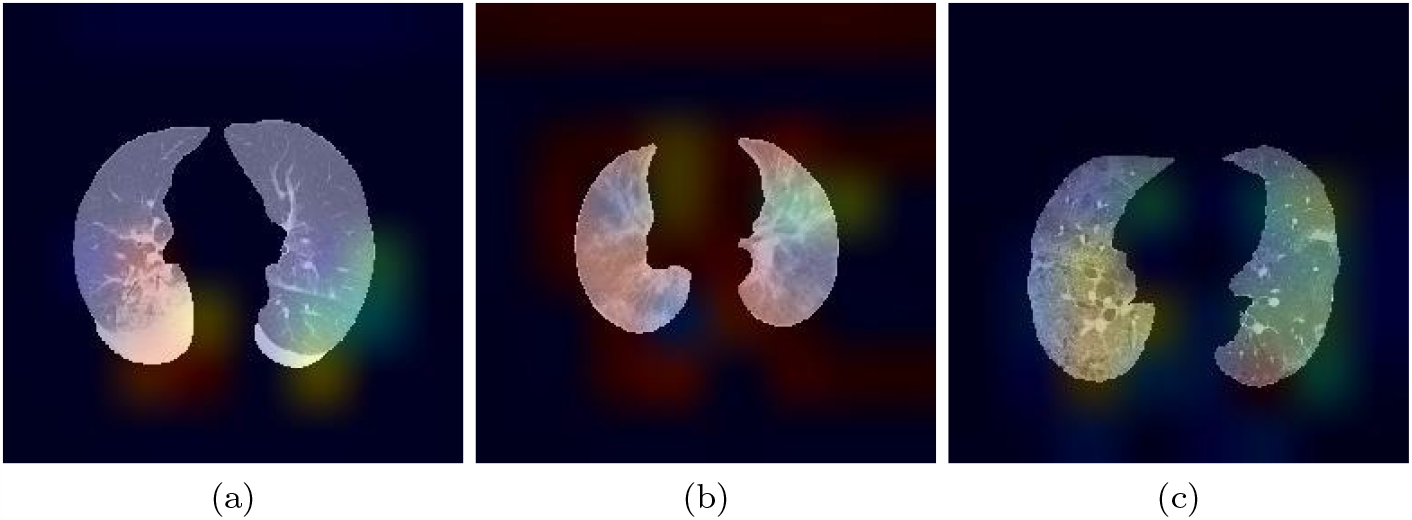
Sample images for the heat map of COVID-19 and other pneumonia images. **(a)** is heat map of Bacterial Pnumonia. **(b)** is heat map of COVID-19.**(c)** is heat map of Viral Pnumonia.

## 4 Discussion and Concluding Remarks

This is the first study to use quantum Fourier transform technique in computed-aided diagnosis of COVID-19 pneumonia. Rapid diagnosis has a key role in the control of infectious diseases and current pandemic of COVID-19. Due to the heavy workload of a large number of infected patients and healthcare professionals, it is predicted that the artificial intelligence-based computer-aided system can speed up the diagnostic process. These systems can enable effective diagnosis of disease in a shorter period of time, as in the case of lack and inability of radiologists during a pandemic crisis. It can also reduce the need for human surveillance and identify details invisible to the human eye.

Patients undergoing Computed Tomography (CT) scan are given a high radiation dose to obtain high-quality images. On the other hand, noise and artifacts occur in CT images when the radiation dose is reduced. This directly affects diagnostic performance. As a solution to this problem, we use a CNN-based network that suppresses noise to remove interference from low-dose CT (LDCT) images. In this study, we proposed a CNN-based method to distinguish COVID-19 pneumonia from other types of pneumonia using low-dose CT images to reduce the radiation used in CT. This method is aimed to increase the diagnostic performance of LDCT image, reduce interobserver and intraobserver variability, decrease the number of false-negative and false-positive results, reduce time consumed in diagnosis, and achieve standardization in evaluating and reporting radiological images. The accuracy, precision, sensitivity, specificity, and F1-score showed high performance of the proposed technique.

Ozcan[47] used the transfer learning approach with 3 different pre-trained GoogleNet, ResNet18, and ResNet50 CNN models to diagnose COVID-19 (131), viral pneumonia (148), bacterial pneumonia (242), and normal (200) cases from chest x-ray data. They have adopted the grid search method for hyperparameter optimization. They achieved the highest performance on the test set (39 COVID-19, 73 bacterial pneumonia, 60 viral pneumonia, and 44 normal). The results of the study are 97.5%accuracy, 95.9% precision, 97.2% sensitivity, 97.9% specificity and 96.6% F1-score. Loey et al.[48] proposed a system for COVID-19 diagnosis using the Generative Adversarial Network(GAN) and pretrained CNN models with deep transfer learning method. Their work is based on GoogleNet, ResNet18, and AlexNet. Since the number of chest x-ray images for COVID-19 is low, their GAN was used to generate more samples. Their accuracy, precision, sensitivity, and F1-score Performance results on the test dataset(COVID-19 (9), Viral Pneumonia (9), Bacterial pneumonia (9), and Normal (9)) among the dataset with a total of 306 images were 80.5%, 80.5%, 84.2%, and 82.4%, respectively. Khan et al.[26] proposed a deep CNN architecture called CoroNet for diagnosing patients infected with COVID-19 from chest X-ray radiographs. In their study, they used a total of 1251 images of 284 COVID-19, 327 bacterial pneumonia, 330 viral pneumonia, and 310 normal patients. The dataset is divided by 80% and 20% for the training and test set, respectively. They used a 4-fold cross-validation approach to evaluate the performance of the 4-class model. The overall performance of the proposed system achieved 89.6% accuracy, 90.0% precision, 89.9% sensitivity, 96.4% specificity, and 89.8% F1-score. Farooq and Hafeez [49] presented a deep learning scheme with a pre-trained ResNet-50 network to detect COVID-19 infected patients called COVID-ResNet. The data set includes a total of 68 COVID-19, 1203 normal, 931 bacterial pneumonia, and 660 viral pneumonia cases. The overall performance of the proposed system achieved 95.0%, accuracy 96.8% precision 96.8% sensitivity, and 96.9% F1-score, respectively. Our study features the first CNN-based application to use 4-class CT datasets (COVID-19, Viral Pneumonia, Bacterial Pneumonia, and Normal) for the differential diagnosis of COVID-19 and other types of pneumonia. Therefore, for comparison of our results with the current literature, we analyzed the studies in which 4-class chest x-ray datasets are used. The comparison of datasets and performance results with other studies are given in Table 5 and Table 6, respectively.

**Table 5.** Comparison of dataset size between our study and other studies.

| Study | Type of Images | COVID-19 | Viral Pneumonia | Bacterial Penumonia | Normal | Total |
| --- | --- | --- | --- | --- | --- | --- |
| Ozcan[47] | X-ray | 131 | 148 | 242 | 200 | 721 |
| Loey et al.[48] | X-ray | 69 | 79 | 79 | 79 | 306 |
| Khan et al.[26] | X-ray | 284 | 327 | 330 | 310 | 1251 |
| Farooq and Hafeez [49] | X-ray | 68 | 660 | 931 | 1203 | 2862 |
| **Our Study** | **CT** | **4387** | **695** | **1157** | **2635** | **7717** |

**Table 6.** Comparison of performance results between our study and other studies.

| Study | Accuracy(%) | Classes | Precision(%) | Sensitivity(%) | Specificity(%) | F1(%) | Test Size |
| --- | --- | --- | --- | --- | --- | --- | --- |
| Ozcan[47] | 97.6 | Bacterial Pneumonia | 100.0 | 97.3 | na | 98.6 | 73 |
|  |  | COVID-19 | 97.5 | 100.0 | na | 98.7 | 39 |
|  |  | Viral Pneumonia | 97.6 | 93.2 | na | 95.3 | 60 |
|  |  | Normal | 95.2 | 100.0 | na | 97.5 | 44 |
|  |  | Overall | 95.9 | 97.2 | 97.9 | 96.6 | 216 |
| Loey et al.[48] | 80.5 | Bacterial Pneumonia | 88.9 | 70.0 | na | 78.38 | 9 |
|  |  | COVID-19 | 100.0 | 100.0 | na | 100.0 | 9 |
|  |  | Viral Pneumonia | 77.8 | 66.7 | na | 71.8 | 9 |
|  |  | Normal | 55.6 | 100.0 | na | 71.4 | 9 |
|  |  | Overall | 80.5 | 84.2 | na | 82.3 | 36 |
| Khan et al.[26] | 89.6 | Bacterial Pneumonia | 86.85 | 95.9 | 95.0 | 86.3 | na |
|  |  | COVID-19 | 93.17 | 98.25 | 97.9 | 95.61 | na |
|  |  | Viral Pneumonia | 84.1 | 82.1 | 94.8 | 83.1 | na |
|  |  | Normal | 95.25 | 93.5 | 98.1 | 94.3 | na |
|  |  | Overall | 90.0 | 89.92 | 96.4 | 89.8 | na |
| Farooq and Hafeez[49] | 95.0 | Bacterial Pneumonia | 95.6 | 97.1 | na | 96.3 | 246 |
|  |  | COVID-19 | 100.0 | 100.0 | na | 100.0 | 8 |
|  |  | Viral Pneumonia | 92.7 | 93.9 | na | 93.3 | 234 |
|  |  | Normal | 99.1 | 96.5 | na | 97.8 | 149 |
|  |  | Overall | 96.8 | 96.8 | na | 96.9 | 637 |
| **Our Study** | **99.5** | Bacterial Pneumoina | 99.6 | 98.7 | 99.9 | 99.1 | 232 |
|  |  | COVID-19 | 99.4 | 99.7 | 99.1 | 99.5 | 646 |
|  |  | Viral Pneumonia | 97.9 | 97.9 | 99.8 | 97.9 | 139 |
|  |  | Normal | 100.0 | 100.0 | 100.0 | 100.0 | 527 |
|  |  | Overall | **99.2** | **99.0** | **99.7** | **99.1** | **1544** |

The performance of a model is measured by its ability to generalize over test data. The data set we used for training in our study is 4-5 times larger than the datasets used in other studies. In addition, their markable point is that the generalization ability of the model is tested on a data set that is approximately 3-4 times the dataset used in other studies. The results obtained according to the performance criteria in Table 6 show that our model is superior to other models. Since any quantum state is moved into phase space with QFT, all possible situations are easily recognized and calculated simultaneously with the effect of the superposition state, and semantically valuable information is extracted. Therefore, the use of quantum Fourier transform in feature extraction makes a great contribution.

## Data Availability

The data set used in this study was collected from Canakkale Onsekiz Mart University Research Hospital with ethical permissions.

## Funding

No payments or services have been received from a third party.

## Conflict of interest

The authors declare that they have no conflict of interest.

## Notes

### Competing Interest Statement

The authors have declared no competing interest.

### Author Declarations

2011-KAEk-27/2020-E.2000060082 number of project has been given ethical committee approval by Clinical Research Ethics Committee of Canakkale Onsekiz Mart University of Republic of Turkey

